# Predictive Value of Lipoprotein(a) for Stroke Recurrence Risk in Embolic Stroke Patients with Different Pathogenesis

**DOI:** 10.1101/2023.09.27.23296262

**Authors:** Xiwa Hao, Aoming Jin, Jing Xue, Aichun Cheng, Zhiyuan Feng, Jinxi Lin, Hao Li, Xia Meng, Baojun Wang, Jie Xu, Yongjun Wang

## Abstract

**Background:** Acute embolic stroke has a high risk of recurrence, including artery-to-artery embolic, cardioembolism, and embolic stroke of undetermined source. Whether lipoprotein(a) contributes to stroke recurrence risk in embolic stroke patients remains unknown.

**Methods:** This prospective cohort biomarker sub-study included 8,076 patients with ischemic stroke or transient ischemic attack from the Third China National Stroke Registry who had measurements of plasma Lp(a) and magnetic resonance imaging sequences and were followed up for 1 year. Cutoffs were set at the 50 mg/dL for Lp(a). Acute embolic stroke was caused by brain embolism from potential embolic sources, which show multiple acute infarctions rather than lacunar infarct by magnetic resonance imaging. Multivariable-adjusted hazard ratio (HR) were calculated using Cox proportional-hazards models for each category to investigate the associations of Lp(a) with stroke recurrence within 1 year.

**Results:** Among the 8,076 patients, the median (interquartile range) age was 63 (55–70) years; 5,627 males (69.7%). In the multiple acute infarctions group, patients with elevated lipoprotein(a) levels had a higher risk of ischemic stroke recurrence than those with non-elevated lipoprotein(a) levels (13.7% vs. 10.5%, hazard ratio, 1.30; 95% confidence interval, 1.02–1.66)after adjustment for potential confounders at one-year follow-up. This association was not observed in the single acute infarction group (8.9% vs. 7.2%, hazard ratio, 1.25; 95% confidence interval, 0.91–1.70). Based on the embolism mechanism in patients with multiple acute infarctions, the stroke recurrence risk significantly increased in patients with elevated lipoprotein(a) levels (14.0% vs. 8.9%; hazard ratio, 1.75; 95% confidence interval, 1.23–2.48) compared with those with non-elevated lipoprotein(a) levels in the embolic stroke of undetermined source group, but not in the artery-to-artery embolism group (13.7% vs. 12.6%; hazard ratio, 0.98; 95% confidence interval, 0.68–1.41) and cardioembolism group (13.5% vs. 9.7%; hazard ratio, 1.50; 95% confidence interval, 0.57–3.97).

**Conclusions:** Elevated lipoprotein(a) levels were significantly associated with ischemic stroke recurrence risk in patients with multiple acute infarctions, especially in patients with embolic stroke of undetermined source. Lipoprotein(a) may be a new therapeutic target for these patients in the future.

**Clinical Perspective:** *What is new?:*

- Elevated lipoprotein(a) levels were associated with risk of stroke recurrence in patients with multiple acute infarctions, especially in patients with embolic stroke of undetermined source.

*What are the clinical implications?:*

- With the coming era of drugs specifically designed to lower lipoprotein(a) levels, these results indicate that lipoprotein(a) may be a new therapeutic target for embolic stroke of undetermined source patients in the future.
- Further study is needed to confirm an association of elevated lipoprotein(a) levels and stroke recurrence risk in Embolic Stroke Patients.

## Introduction

Acute infarct patterns are an important factor to evaluate the prognosis of patients with ischemic stroke (IS). Substantial evidence from clinical studies has shown that patients with multiple acute infarctions (MAIs) have a much higher risk of stroke recurrence than those with single acute infarction (SAI) or no acute infarction, which indicates that MAIs are important imaging markers to predict stroke recurrence.^1–3^ Multiple infarctions indicate a mechanism of embolism,^4^ including artery-to-artery (A-to-A) embolism,^5^ cardioembolism (CE),^6^ and embolic stroke of undetermined source (ESUS).^7^

Elevated lipoprotein(a) [Lp(a)] is an independent risk factor for IS and can predict the risk of stroke recurrence.^8–11^ Lp(a) potentially contributes to cardiovascular disease through the proatherogenic effects of its LDL-like moiety, proinflammatory effects of its oxidized phospholipid content, and prothrombotic effects through its inactive, plasminogen-like protease domain on apolipoprotein(a).^12–14^ Previous studies have reported that the prothrombotic and proinflammatory effects of Lp(a) have been suggested to promote plaque destabilization, leading to plaque rupture and embolism events.^15, 16^ Therefore, we hypothesized that elevated Lp(a) levels may be an important biomarker for the pathogenesis of A-to-A embolism. However, the association between Lp(a) and other types of embolic stroke, especially cardiogenic stroke and ESUS, remains unclear.

We aimed to investigate patients with IS and transient ischemic attack (TIA), by differentiating SAI and MAI according to infarction patterns. First, the association between Lp(a) and the risk of stroke recurrence was analyzed according to SAI and MAIs. Furthermore, we divided the patients with MAIs into A-to-A embolism, CE, and ESUS groups to explore the association between the Lp(a) level and stroke recurrence.

## Methods

### Study population

This study derived data from the biomarker sub-study of the Third China National Stroke Registry (CNSR-III). Details about the rationale, design, and results of this study have been published elsewhere.^17^ In brief, the sub-study consisted of 11,261 individuals recruited among patients with acute IS or TIA in 171 hospitals. Participants aged >18 years were enrolled in this study between August 2015 and March 2018. All patients in this sub-study were asked to undergo magnetic resonance imaging (MRI) examinations and complete blood samples during hospitalization. Patients with the following MRI sequences were included in the current analysis: T1-weighted; T2-weighted; and diffusion-weighted imaging (DWI). Patients without baseline MRI sequences and Lp(a) levels were excluded. The protocol and data collection methods of the CNSR-III were approved by the ethics committee of Beijing Tiantan Hospital and other participating hospitals. Written informed consent was obtained from each participant or their legally authorized representative before entering the study.

### Image analysis and interpretation

All patient image data during hospitalization were collected in the DICOM format on discs and analyzed by the image research center in Beijing Tiantan Hospital based on centralized forms. All images and Trial of Org 10172 in Acute Stroke Treatment (TOAST) classification were visually evaluated by two experienced readers, and discrepancies were resolved by a third reader. Acute IS and infarction patterns (SAI and MAIs) were diagnosed with hyperintense lesions on DWI sequences according to infarction number; the presence of SAI was defined as uninterrupted lesions visible in contiguous territories, while the presence of MAIs was defined as more than one lesion that was topographically distinct (separated in space or discrete on contiguous slices).^2, 18^ TIA was defined as focal brain ischemia with resolution of symptoms within 24 hours after onset. Trained assessors blinded to the clinical data of the patients independently evaluated the infarction patterns. A-to-A embolic stroke was defined as a large-artery atherosclerosis stroke according to the TOAST classification and MAIs seen at the cortical and subcortical regions on DWI sequences, but not involving border-zone areas, except for hemodynamic deficiency attributable to concurrent stenoses.^19^ Criteria for diagnosis of ESUS: (1) stroke detected by CT or MRI that is not lacunar; (2) absence of extracranial or intracranial atherosclerosis causing ≥50% luminal stenosis in arteries supplying the area of ischemia; (3) no major risk cardioembolic source of embolism; and (4) no other specific cause of stroke identified (e.g., arteritis, dissection, migraine/vasospasm, drug misuse).^20^

### Laboratory methods

EDTA fasting blood samples were collected from 171 study sites within 24 hours of admission. All blood samples were transported to the center laboratory at Beijing Tiantan Hospital in a maintained cold chain and stored at -80℃ until testing. Lp(a) concentrations were determined using enzyme-linked immunosorbent assay (ELISA) kits (Mercodia AB, Uppsala, Sweden). The Mercodia Lp(a) ELISA is based on the direct sandwich technique, in which two monoclonal antibodies are directed against separate antigenic determinants on the Apo(a) molecule. This is a well-validated assay with good reproducibility (coefficient of variation, 5.52–6.98%), which yields robust results and is generally insensitive to apolipoprotein(a) size heterogeneity.

### Patient follow-up and outcome evaluation

Patients were followed up clinically by face-to-face interviews at discharge and at three months, and trained research coordinators contacted the patients by phone at six months and one year. The details of the patient follow-up have been previously described.^21^ The primary outcome was IS recurrence (time-to-first) within one year. IS was defined as an acute focal infarction of the brain or retina with one of the following: sudden onset of a new focal neurological deficit lasting less than 24 hours with clinical or imaging evidence of infarction, or rapid worsening of an existing focal neurological deficit lasting 24 hours or more, with imaging evidence of new ischemic changes clearly distinct from the index ischemic event.

### Statistical analysis

Baseline patient characteristics were compared between patients with Lp(a) ≥50 mg/dL and <50 mg/dL in SAI and MAIs based on MRI-DWI sequences. Continuous variables are expressed as medians with interquartile ranges, and categorical variables are presented as frequencies and percentages. Baseline variables were compared between groups using the χ^2^ test or Fisher’s exact test for categorical variables and one-way ANOVA or Kruskal– Wallis test for continuous variables with normal or skewed distribution, respectively.

Patients were divided into several groups according to different acute infarction patterns and Lp(a) levels (non-elevated Lp(a) level <50 mg/dL and elevated Lp(a) level ≥50 mg/dL, as proposed by the European Atherosclerosis Society and American Heart Association and American College of Cardiology^22^). The percentage of patients with primary outcome events within one year of follow-up was estimated using Kaplan–Meier survival analysis, and the associations of baseline Lp(a) with stroke recurrence risk were investigated using Cox proportional-hazards models for each group within one year. Covariates correlated with Lp(a) were identified based on a baseline variable comparison between the two Lp(a) categories. Moreover, the association between the variables and outcome was assessed based on clinical knowledge. According to the definition of confounders that correlated with the exposure and outcome, covariates, including age, sex, BMI, history of stroke, history of coronary heart disease, TOAST subtype, eGFR, HDL-C, and LDL-Cc, were included in the adjusted model. Multivariable adjusted hazard ratios (HRs) with their corresponding 95% confidence intervals (CI) were calculated. A two-sided P value of <0.05 was considered statistically significant. SAS software (version 9.4; SAS Institute, Inc., Cary, NC) was used for all the statistical analyses.

## Results

### Patient demographics and baseline characteristics

From August 2015 to March 2018, 11,261 patients with IS or TIA were recruited for the biomarker sub-study of CNSR-III. After excluding 583 patients without baseline Lp(a) levels available, 1,443 patients without T1-weighted imaging, T2-weighted imaging, and DWI, and 1,159 patients without hyperintense lesions on DWI, 8,076 patients (71.7%) remained with all the required sequences and were included in the current analysis (Figure 1). For all participants, the median of the 25th and 75th percentiles for the follow-up duration was 365 days (360 and 365 days, respectively).

**Figure 1.**
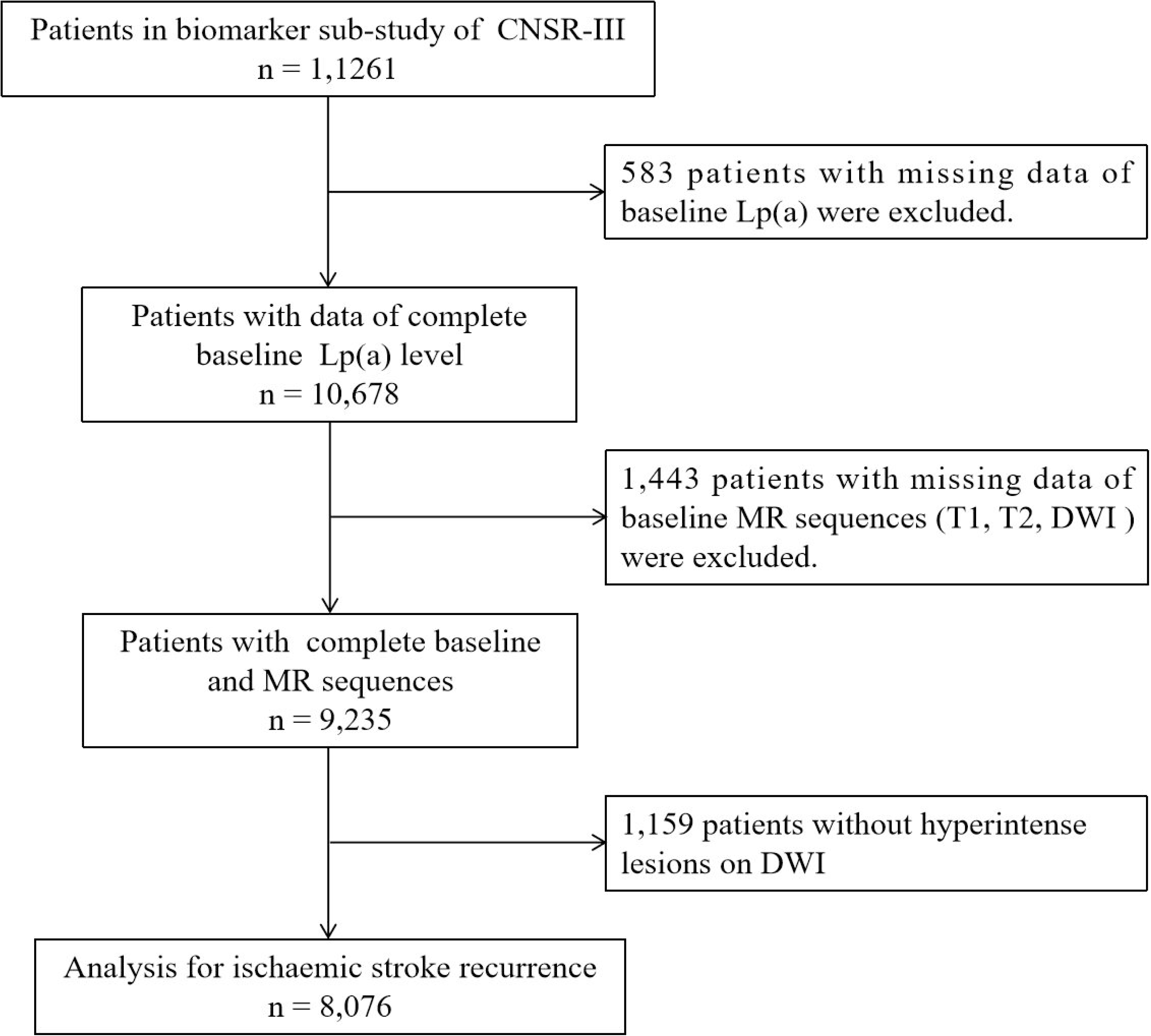
Flow diagram of participants in this study.

Among the 8,076 patients, the median (Interquartile range,IQR) age was 63 (55–70) years, and 5,627 (69.7%) were males. Within one year, 749 (9.3%) patients experienced at least one stroke recurrence. A total of 4,122 patients (51.0%) had MAIs, and 3,954 patients (49.0%) had SAI. Table 1 shows the demographic and clinical characteristics of the patients with different infarction patterns (SAI and MAIs) and baseline Lp(a) levels (≥50 mg/dL and <50 mg/dL). Patients with elevated Lp(a) levels and MAIs had a higher proportion of previous history of stroke and coronary heart disease and were less likely to have known atrial fibrillation than those with non-elevated Lp(a) levels and MAIs. However, patients with elevated Lp(a) levels and SAI had higher inflammation levels (IL-6 and hs-CRP) than those with non-elevated Lp(a) levels and SAI.

**Table 1.** Baseline characteristics of patients stratified according to Lp(a) levels and acute infarction patterns.

|  | SAI |  |  |  | MAI |  |  |  |
| --- | --- | --- | --- | --- | --- | --- | --- | --- |
|  | Total patients n =<br>3954 | Non-elevated Lp(a)<br>level n = 3347 | Elevated Lp(a) level<br>n = 607 | p | Total patients n =<br>4122 | Non-elevated Lp(a)<br>level n = 3471 | Elevated Lp(a) level<br>n = 651 | p |
| Age, years, median (IQR) | 62 (54–69) | 62 (54–70) | 62 (55–68) | 0.83 | 63 (55–71) | 63 (55–71) | 63 (55–71) | 0.59 |
| Male, n (%) | 2,697 (68.2) | 2,304 (68.8) | 393 (64.7) | 0.05 | 2,930 (71.1) | 2,479 (71.4) | 451 (69.3) | 0.27 |
| Current smoker, n (%) | 1,279 (32.4) | 1,091 (32.6) | 188 (31.0) | 0.43 | 1361 (33.0) | 1,162 (33.5) | 199 (30.6) | 0.15 |
| BMI, kg/m<sup>2</sup>, median (IQR) | 24.49 (22.76–26.57) | 24.54 (22.85–26.60) | 24.24 (22.32–26.04) | 0.02 | 24.42 (22.49–26.57) | 24.47 (22.49–26.57) | 24.22 (22.31–26.57) | 0.21 |
| **Type of stroke, n (%)** |  |  |  | 0.46 |  |  |  | 0.34 |
| Ischemic stroke, n (%) | 3,913 (99.0) | 3,314 (99.0) | 599 (98.7) |  | 4,083 (99.1) | 3,436 (99.0) | 647 (99.4) |  |
| TIA, n (%) | 41 (1.0) | 33 (1.0) | 8 (1.3) |  | 39 (0.9) | 35 (1.0) | 4 (0.6) |  |
| **Diseases history, n (%)** |  |  |  |  |  |  |  |  |
| Hypertension | 2,555 (64.6) | 2,175 (65.0) | 380 (62.6) | 0.26 | 2,538 (61.6) | 2,139 (61.6) | 399 (61.3) | 0.87 |
| Diabetes mellitus | 963 (24.4) | 821 (24.5) | 142 (23.4) | 0.55 | 969 (23.5) | 806 (23.2) | 163 (25.0) | 0.32 |
| Stroke | 816 (20.6) | 691 (20.7) | 125 (20.6) | 0.98 | 973 (23.6) | 798 (23.0) | 175 (26.9) | 0.03 |
| Coronary heart disease | 327 (8.3) | 282 (8.4) | 45 (7.4) | 0.40 | 484 (11.7) | 393 (11.3) | 91 (14.0) | 0.05 |
| Atrial fibrillation | 135 (3.4) | 116 (3.5) | 19 (3.1) | 0.68 | 468 (11.4) | 414 (11.9) | 54 (8.3) | 0.01 |
| **TOAST classification, n (%)** |  |  |  | 0.24 |  |  |  | 0.002 |
| Large-artery<br>atherosclerosis | 486 (12.3) | 404 (12.1) | 82 (13.5) |  | 1,692 (41.1) | 1,399 (40.3) | 293 (45.0) |  |
| Cardioembolic | 172 (4.4) | 147 (4.4) | 25 (4.1) |  | 345 (8.4) | 308 (8.9) | 37 (5.7) |  |
| Microangiopathic | 2,105 (53.2) | 1,795 (53.6) | 310 (51.1) |  | - | - | - |  |
| Other cause | 39 (1.0) | 37 (1.1) | 2 (0.3) |  | 54 (1.3) | 40 (1.2) | 14 (2.2) |  |
| Unknown cause | 1,152 (29.1) | 964 (28.8) | 188 (31.0) |  | 2,031 (49.3) | 1,724 (49.7) | 307 (47.2) |  |

**Table 1. Baseline characteristics of patients stratified according to Lp(a) levels and acute infarction patterns (continued).**
|  | SAI |  |  |  | MAI |  |  |  |
| --- | --- | --- | --- | --- | --- | --- | --- | --- |
|  | Total patients n = 3954 | Non-elevated Lp(a)<br>level n = 3347 | Elevated Lp(a)<br>level n = 607 | p | Total patients n<br>= 4122 | Non-elevated Lp(a)<br>level n = 3471 | Elevated Lp(a)<br>level n = 651 | p |
| **Discharge medication, n (%)** |  |  |  |  |  |  |  |  |
| Antiplatelet | 3,758 (95.1) | 3,181 (95.1) | 577 (95.2) | 0.93 | 3,676 (89.5) | 3,083 (89.2) | 593 (91.4) | 0.09 |
| Anticoagulant | 53 (1.3) | 45 (1.4) | 8 (1.3) | 0.96 | 204 (5.0) | 181 (5.2) | 23 (3.5) | 0.07 |
| Anti-hypertension | 2,176 (55.1) | 1,859 (55.6) | 317 (52.3) | 0.14 | 1,892 (46.1) | 1,601 (46.3) | 291 (44.8) | 0.49 |
| Statin | 3,721 (94.1) | 3,153 (94.2) | 568 (93.6) | 0.55 | 3,790 (92.0) | 3,179 (91.6) | 611 (93.9) | 0.05 |
| Antidiabetics | 1,021 (25.9) | 868 (26.0) | 153 (25.3) | 0.71 | 994 (24.2) | 837 (24.2) | 157 (24.2) | 0.99 |
| **Other** |  |  |  |  |  |  |  |  |
| eGFR (mL/min/1.73 m<sup>2</sup>) | 93.76 (83.25–102.42) | 93.85 (83.59–<br>102.49) | 93.59 (81.97–<br>101.30) | 0.25 | 92.21 (79.43–<br>101.29) | 92.66 (80.19–<br>101.68) | 89.84 (75.52–<br>98.86) | <0.001 |
| FPG (mmol/L), median (IQR) | 5.52 (4.90–7.10) | 5.50 (4.90–7.10) | 5.61 (4.98–7.07) | 0.29 | 5.60 (4.90–6.94) | 5.60 (4.93–7.00) | 5.50 (4.84–6.59) | 0.07 |
| SBP at admission, mmHg, median (IQR) | 150.50 (137.50–<br>166.50) | 150.50 (138.00–<br>166.50) | 150.00 (136.50–<br>166.50) | 0.70 | 148.50 (134.50–<br>164.00) | 148.50 (134.50–<br>164.00) | 147.50 (134.00–<br>162.50) | 0.29 |
| **Lipid profile** |  |  |  |  |  |  |  |  |
| HDL-C (mmol/L), median (IQR) | 0.93 (0.78–1.11) | 0.92 (0.77–1.10) | 0.96 (0.79–1.18) | 0.003 | 0.93 (0.77–1.11) | 0.92 (0.76–1.10) | 0.95 (0.78–1.16) | 0.01 |
| LDL-C (mmol/L), median (IQR) | 2.35 (1.75–3.02) | 2.29 (1.73–2.97) | 2.62 (2.01–3.35) | <0.001 | 2.32 (1.74–2.98) | 2.27 (1.70–2.92) | 2.60 (2.00–3.44) | <0.001 |
| LDL-Cc (mg/dL), median (IQR) | 82.52 (60.34–108.38) | 83.57 (61.64–<br>109.09) | 75.66 (50.53–<br>104.52) | <0.001 | 81.28 (59.14–<br>107.48) | 82.09 (60.29–<br>107.39) | 74.63 (50.02–<br>108.26) | <0.001 |
| Lp(a) (mg/dL), median (IQR) | 17.86 (8.76–34.35) | 14.84 (7.61–24.82) | 74.66 (60.50–<br>103.85) | <0.001 | 18.36 (9.12–<br>36.64) | 15.21 (8.00–26.12) | 76.23 (60.01–<br>104.52) | <0.001 |
| **Inflammatory biomarkers** |  |  |  |  |  |  |  |  |
| hs-CRP (mg/dL), median (IQR) | 1.44 (0.74–3.67) | 1.41 (0.74–3.54) | 1.59 (0.76–4.31) | 0.07 | 2.34 (0.96–6.22) | 2.34 (0.96–6.13) | 2.38 (0.94–6.88) | 0.67 |
| IL-6 (ng/L), median (IQR) | 2.38 (1.51–4.15) | 2.33 (1.51–4.07) | 2.57 (1.58–4.81) | 0.01 | 3.25 (1.93–6.41) | 3.22 (1.91–6.31) | 3.52 (2.01–6.97) | 0.10 |

Lp(a), lipoprotein(a); SAI, single acute infarction; MAI, multiple acute infarction; IQR, interquartile range; BMI, body mass index; TIA, transient ischemic attack; TOAST, Trial of Org 10172 in Acute Stroke Treatment; eGFR, estimated glomerular filtration rate; FPG, fasting plasma glucose; SBP, systolic blood pressure; HDL-C; high-density lipoprotein cholesterol; LDL-C, low-density lipoprotein cholesterol; hs-CRP, high-sensitivity C-reactive protein; IL-6, interleukin-6.

### Effect of Lp(a) on stroke recurrence according to acute infarction patterns

As shown in Figure 2A, Kaplan–Meier analysis with the log-rank test showed that participants with elevated Lp(a) levels and MAIs had a significantly higher cumulative recurrence rate than those with non-elevated Lp(a) levels and MAIs (p =.0171). Conversely, the cumulative recurrence rate was not significantly different between the patients with SAI groups (Figure 2B). After multivariable adjustment, elevated Lp(a) levels remained significantly associated with stroke recurrence risk (13.7% vs 10.5%, HR, 1.30; 95% CI, 1.02–1.66; p =.03) in patients with MAIs. This association was not observed in patients with SAI (8.9% vs. 7.2%, HR, 1.25; 95% CI, 0.91–1.70; p =.44) (Table 2).

**Figure 2.**
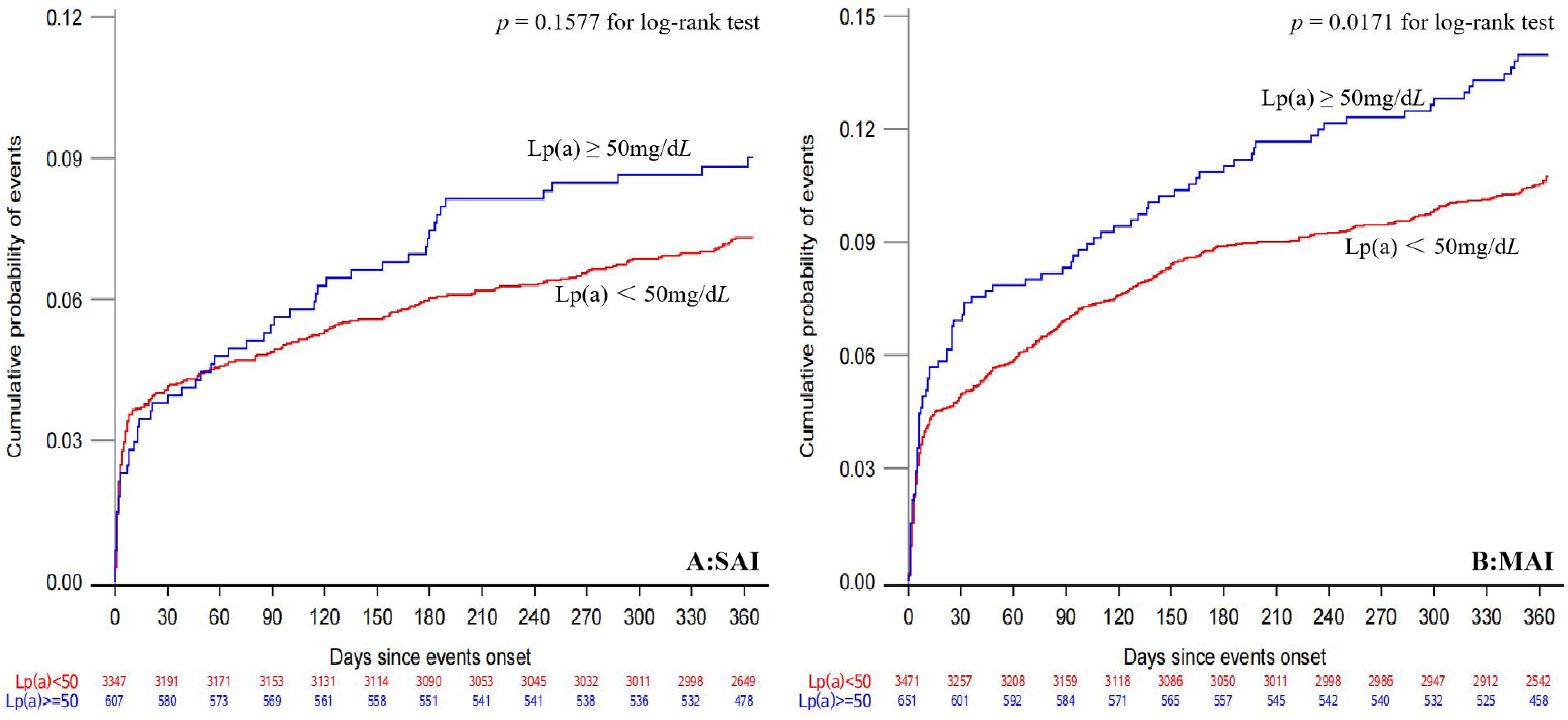
Kaplan–Meier curves of cumulative risk for stroke recurrence by Lp(a) level and acute infarction. A. Kaplan–Meier curves showing the time to event for the primary outcome according to baseline Lp(a) level (non-elevated Lp(a) level <50 mg/dL and elevated Lp(a) level ≥50 mg/dL) in patients with MAIs B. Kaplan– Meier curves showing the time to event for the primary outcome according to baseline Lp(a) level (non-elevated Lp(a) level <50 mg/dL and elevated Lp(a) level ≥50 mg/dL) in patients with SAI.

**Table 2.** Risk of clinical events by Lp(a) level and acute infarction patterns.

| Outcome | Group | No | Events at 1 year, n (%) | Model 1<sup>a</sup> |  | Model 2<sup>b</sup> |  |
| --- | --- | --- | --- | --- | --- | --- | --- |
|  |  |  |  | Adjusted HR (95% CI) | *P* | Adjusted HR (95% CI) | *P* |
| Stroke recurrence | Non-elevated Lp(a) level with SAI | 3347 | 242 (7.2) | Ref | 0.17 | Ref | 0.16 |
|  | Elevated Lp(a) level with SAI | 607 | 54 (8.9) | 1.23 (0.91–1.65) |  | 1.25 (0.91–1.70) |  |
|  | Non-elevated Lp(a) level with MAI | 3471 | 364 (10.5) | Ref | 0.02 | Ref | 0.03 |
|  | Elevated Lp(a) level with MAI | 651 | 89 (13.7) | 1.31 (1.04–1.65) |  | 1.30 (1.02–1.66) |  |

<sup>a</sup>Adjusted for age and sex.
<sup>b</sup>Adjusted for age, sex, BMI, history of stroke, history of coronary heart disease, TOAST subtype, GFR, HDL-C, and LDL-Cc.
Non-elevated Lp(a) level <50 mg/dL; Elevated Lp(a) level ≥50 mg/dL.
Lp(a), lipoprotein(a); SAI, single acute infarction; MAI, multiple acute infarction; CI, confidence interval; HR, hazard ratio.

### Association of Lp(a) with stroke recurrence risk in patients with MAIs according to embolism mechanism

We divided the patients with MAIs into three groups based on the embolism mechanism (A-to-A embolism, CE, and ESUS). In patients with MAIs characterized by ESUS, the stroke recurrence risk significantly increased in patients with elevated Lp(a) levels compared with those with non-elevated Lp(a) levels (14.0% vs. 8.9%; HR, 1.75; 95% CI, 1.23–2.48). This association was not observed in patients with A-to-A embolism and CE (13.7% vs. 12.6%; HR, 0.98; 95% CI, 0.68–1.41) and (13.5% vs. 9.7%; HR, 1.50; 95% CI, 0.57–3.97) (Figure 3).

**Figure 3.**
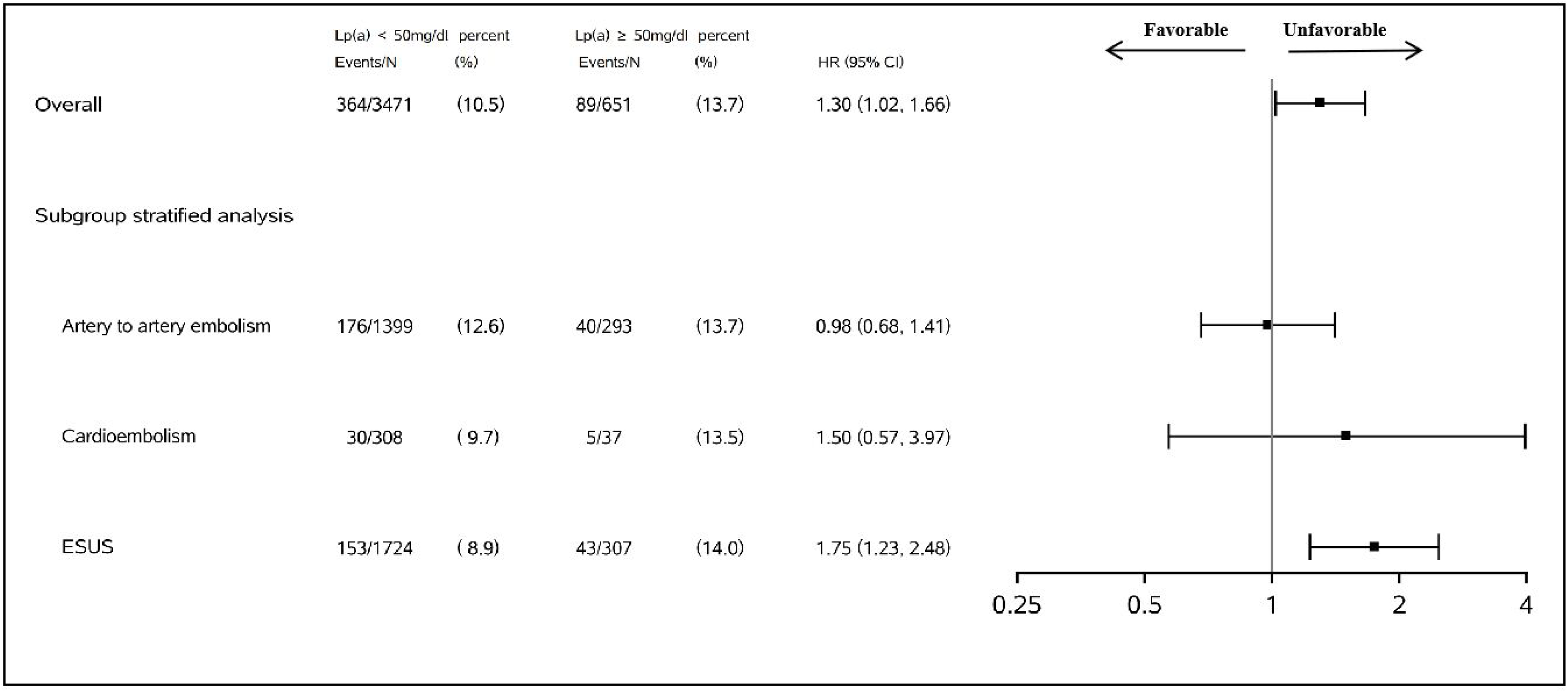
Stroke recurrence association with Lp(a) according to embolism mechanism in patients with multiple acute infarctions. Adjusted variables: age, sex, BMI, history of stroke, history of coronary heart disease, TOAST subtype, GFR, HDL-C, and LDL-Cc. ESUS, embolic stroke of undetermined source.

## Discussion

In this study, we showed that elevated Lp(a) levels were associated with the risk of stroke recurrence in patients with MAIs, and this finding was not observed in patients with SAI. In grouping patients with MAIs according to pathogenesis, we found that Lp(a) was an independent risk factor for stroke recurrence in ESUS patients rather than in A-to-A embolic stroke. In addition, we found that elevated Lp(a) levels were associated with a higher risk of stroke recurrence than non-elevated Lp(a) levels in patients with CE; however, the difference was not statistically significant.

Baseline imaging features, especially different MRI sequences, could provide vital information on the underlying etiology and mechanism of IS. Evidence exists that MAIs detected by DWI had a higher risk of recurrent stroke and vascular events in patients with IS or TIA.^18, 23^ The Clopidogrel in High-Risk Patients With Acute Nondisabling Cerebrovascular Events (CHANCE) randomized clinical trial has demonstrated stroke recurrence occurred in 15 (10.1%) and 25 (18.8%) of patients with MAIs administered clopidogrel plus aspirin and placebo plus aspirin, respectively; and 24 (8.9%) and 24 (8.5%) of patients with SAI administered clopidogrel plus aspirin and placebo plus aspirin at three-month follow-up.^3^ Previous studies indicated that MAIs are usually related to large-artery atherosclerosis (LAA), cardioembolism, and undetermined cause stroke according to the TOAST classification.^24^ In addition, the pathogenesis of MAIs is likely caused by the embolism from heart or major extracranial or intracranial artery (A-to-A embolism, CE, and ESUS).^20, 25, 26^ Our study showed that the proportion of stroke recurrence significantly increased in patients with elevated Lp(a) levels and MAIs. Specifically, the stroke recurrence risk significantly increased in patients with elevated Lp(a) levels compared with those with non-elevated Lp(a) levels (13.67% vs. 10.49%; adjusted HR, 1.30; 95% CI, 1.02– 1.66). A small case-control study showed that a high serum Lp(a) level is an independent risk factor for stroke patients with MAIs diagnosed by computed tomography.^27^ In our study, we observed a similar association. Therefore, elevated Lp(a) levels may be useful as a risk assessment biomarker for recurrent stroke in patients with MAIs.

Elevated Lp(a) levels can help to identify the underlying causes of IS. A meta-analysis of 45 studies revealed that Lp(a) levels were significantly associated with the risk of the LAA subtype of IS.^8^ Previous studies also showed that elevated Lp(a) levels were independently associated with the risk of stroke or recurrent vascular events, particularly arteriosclerotic disease.^11, 21, 28^ The reasons why a high Lp(a) level may increase the risk of stroke or vascular event recurrence are attributed to proatherogenic, proinflammatory, and prothrombotic effects. Unexpectedly, our study results showed that elevated Lp(a) levels were independently associated with the risk of stroke recurrence in ESUS patients with MAI, but not in those with the LAA subtype. Furthermore, this association was not observed in patients with CE; however, we believe that the small sample size and low rate of stroke recurrence may be the main reasons for the absence of statistical significance. Recent studies found that most strokes recurrent in undetermined cause and CE patients were embolic of undetermined source or cardiac.^29, 30^ A small clinical study showed that the serum Lp(a) level is higher in patients with IS associated with atrial fibrillation and left atrial thrombus formation, suggesting that the plasminogen inhibitory action of Lp(a) may play an important role in left atrial thrombus formation.^31^ Our results indicate that Lp(a) contributes to the risk of stroke recurrence largely through a prothrombotic mechanism.

Given the underlying causes of stroke recurrence in patients with elevated Lp(a) levels and MAIs, lower Lp(a) therapy may reduce the risk of recurrent stroke. The Apo(a) component of Lp(a) is highly homologous to plasminogen but has no fibrinolytic activity. Data suggests that Apo(a) inhibits activation of plasminogen and increases in the expression of PAI-1 to inhibit fibrinolysis and the inactivation of tissue factor pathway inhibitor, which augments factor VII activation and promotes thrombosis.^32^ Interestingly, a recent study found that elevated Lp(a) levels during the acute phase of COVID-19 were strongly associated with venous thromboembolism incidence.^33^ A meta-analysis of fourteen studies, comprising a total of more than 14,000 patients, showed that Lp(a) appeared to be significantly associated with increased risk of venous thromboembolism.^34^ Previous studies further support our results. Our study can support identification of high-risk stroke patients including Lp(a) levels to prevent stroke recurrence in patients with ESUS and CE.

### Limitations

The strengths of our study are that the interpretation of infarct pattern images and TOAST classification for more than 8,000 patients which were performed centrally at the Imaging Center of Beijing Tiantan Hospital. In addition, baseline Lp(a) concentrations were centrally measured by uniformly collected blood samples. Our study also has some limitations. First, imaging examinations were not collected during the follow-up of this study, so the interpretation of infarction patterns and TOAST classification could not be carried out for recurrent stroke events. However, previous studies have shown that the pathogenesis of recurrence events and index events are often the same, especially for LAA and CE patients.^35^ Second, in this study, the sample size of CE patients was small. Therefore, the trend of association between Lp(a) and stroke recurrence in patients with CE needs to be further verified in studies with large sample sizes, especially in Caucasian populations with a higher CE incidence.

## Conclusions

Elevated Lp(a) levels are significantly associated with stroke recurrence risk in patients with MAIs, especially ESUS. These results primarily reinforces the value of Lp(a) testing in stroke patients for more refined risk classification and hence, enhanced disease management.

## Data Availability

Contact the corresponding author via email

## Acknowledgments

We thank the participants and staff of the Third China National Stroke Registry (CNSR-III) study.

## Funding Sources

This study is supported by grants from the National Natural Science Foundation of China (81870905, U20A20358, 82111530203), Chinese Academy of Medical Sciences Innovation Fund for Medical Sciences (2019-I2M-5-029), and the Capital’s Funds for Health Improvement and Research (2020-1-2041), National Key R&D Program of China (2022YFC2502403)

## Conflict of Interest Disclosures

None reported.

## Notes

### Competing Interest Statement

The authors have declared no competing interest.

### Author Declarations

the ethics committee of Beijing Tiantan Hospital and other participating hospitals

